# Identifying contextual and patient-level factors associated with the utilization of the ‘Understand me!’ heart rate-informed communication tool in care for non-communicative patients: A study protocol

**DOI:** 10.1101/2025.11.03.25339425

**Authors:** Maiken E. T. Willadsen, Bjørnar Hassel, Daniel Quintana, Emilie S. M. Kildal

**Affiliations:** Department of Psychology, University of Oslo, Norway; Department of Neurohabilitation, Oslo University Hospital, Oslo, Norway; Department of Clinical Medicine, University of Oslo, Norway; K.G. Jebsen, Centre for Neurodevelopmental Disorder, University of Oslo, Norway; NevSom, Department of Rare Disorders, Oslo University Hospital, Oslo, Norway

## Abstract

**Background:** Some patients with profound intellectual and multiple disabilities (PIMD) have major communication difficulties, making them unable to notify their caregivers when they experience pain or stress. These people have been referred to as minimally communicative patients (MCP). Preliminary studies have found that heart rate (HR) monitoring can be used for understanding potential painful or stressful procedures in MCPs. For such a communication tool to function effectively, it must be actively utilized. However, obstacles to the adoption of this communication tool have been insufficiently explored, particularly regarding the feasibility of HR-informed communication tool in the care of MCPs.

**Objectives:** The study explores contextual and patient-level factors associated with the utilization of the “Understand me!” HR-informed communication tool. Utilization is quantified primarily as hours of device use and secondarily as the caregiver’s response rate to system alarms and the number of registered situations.

**Hypotheses:** *Hypothesis 1:* Patients in larger staffed residential units (≥ 8 staff) will have a lower proportion of alarms answered by caregivers than smaller units (< 8 staff), adjusted for patient function level. *Hypothesis 2:* There will be a positive correlation between the number of distinct caregivers and the proportion of alarms answered by caregivers, adjusting for hours of device exposure and number of alarms. *Hypothesis 3:* Patients classified as immobile will have a higher proportion of alarms answered by caregivers than mobile patients, after accounting for hours of monitoring and number of alarms. *Hypothesis 4:* Greater patient medication burden will be associated with a higher proportion of alarms answered by caregivers, compared with lower medication burden, after adjustment for patient function level and size of residential unit.

**Methods:** Analysis of factors involved in utilization for caregivers of 40 non-communicative patients. Data will be collected during a clinical testing phase of 2-3 months where caregivers will be asked to report contexts for when the app alerted them of a stark increase in HR.

**Discussion:** This study will advance the field of wearable physiological sensor use in patient care by providing insight into various contextual and patient-level factors that may influence the use of assistive devices.

**Trial registration:** The larger clinical trial that this study is part of was registered prospectively at ClinicalTrials.gov (NCT05738278).

## Introduction

Assistive devices are often used in the care of patients with profound intellectual and multiple disabilities (PIMD) to aid communication. Patients with PIMD are heavily dependent on their caregivers (parents, guardians, family members, or healthcare staff) to understand and support them for all aspects of their day (Maes et al., 2007). PIMD is the combination of profound intellectual disability (IQ < 20) and inability to move independently (Matérne & Holmefur, 2022). Due to their communication difficulties, it is challenging to interpret their needs, wishes, and preferences, and they are often misunderstood (Maes et al., 2007). These patients can be referred to as minimally communicative patients (MCP). They are dependent on caregivers being aware of their needs (Bradshaw, 2001; Øderud et al., 2023). Patients with PIMD will typically need the equivalent of 5 – 10 full-time staff per year (Regjeringen, 2009). Due to the increasing need for customized and intensive nursing care, there is a growing interest in technological innovations that can improve communication and engagement for this vulnerable population (Ali & Hassiotis, 2008).

MCPs can experience poor follow-up from caregivers. Children with MCP have been recognised as being at greater risk of experiencing pain, and they may experience pain during daily care activities. In circumstances like this, the inability to communicate pain or stress represents the key problem (Cascella et al., 2019). Heart rate (HR) can be used for understanding potential painful or stressful procedures in MCPs (Kildal et al., 2021; 2023). Consequently, HR-guided interventions may help caregivers adapt everyday care situations to MCPs (Kildal et al., 2021).

MCPs are often in potentially stressful and painful situations, such as physiotherapy, using casts for spastic limbs, when lifted, and when their personal hygiene is being attended to. The most important reason to use such a communication tool is to understand which activities are experienced as painful or unpleasant, to avoid or tailor these for the benefit of the patient (De Boer et al., 2009). Not using an assistive device can increase the likelihood of experiencing excessive caregiver burden (Hwang et al., 2025). Despite caregivers’ intention to enhance the daily experiences of patients and their desire to use such a communication tool, its utilization remain insufficient (Baily et al., 2006). The effectiveness of communication tools are contingent upon its comprehensive integration across the caregivers, requiring consistent engagement from throuhout the care of the patient. Obstacles to the use of communication tools for MCPs are generally poorly studied. The feasibility and sustained use of HR-monitoring tools in residential care settings may depend on institutional factors— particularly the size and organization of the residence. Evidence suggests that smaller residential units, where caregivers have more one-to-one time with each resident, are associated with higher quality of care (Spilsbury et al., 2024). This may be the result of staff members typically knowing the residents better, experiencing less turnover, and being more motivated to adopt individualized monitoring tools such as HR sensors. Moreover, smaller units often have simpler organizational structures, which may impact feasibility. On the other hand, studies have shown that individual caregiver characteristics can moderate technology adoption. For example, Bailey and colleagues (2006) found that the use of communication aids varied among caregivers depending on their understanding of the resident’s non-symbolic communication (e.g., facial expressions, idiosyncratic gestures). Caregivers—especially family members or long-term staff—who already have a sophisticated intuitive understanding of the person’s signals may perceive less need for assistive tools and therefore use them less frequently. Staffing and workload in residential units have been shown as a factor affecting the time available for caregivers to use assistive devices (Noble & Sweeney, 2018). In Norway, MCPs often have two-to-one staffing, and the residential units with the smallest teams have 6 caregivers. Therefore, here we define “larger” residential units as those with at least 8 caregivers.

Patient mobility levels can serve as a significant patient-level factor that can impact caregiver engagement with HR-monitoring tools. Patients with PIMD often experience immobility and require comprehensive mobility support, resulting in caregivers spending considerable time with them. The constant proximity required for care may, in practice, increase opportunities for caregivers to observe HR data and respond to alarms. Research indicates that mobile patients tend to exhibit more frequent behavioral disturbances than immobile patients (Miyamoto & Kurita, 2002). These mobile patients may wander and display aggression—behaviors that are known predictors of increased caregiver resource requirements (Miyamoto & Kurita, 2002), and such behavioral tendencies in intellectually disabled mobile patients can impede caregivers’ ability to respond to alerts from HR monitoring devices.

Medications represent a crucial component in the treatment, management, and prevention of various medical conditions (Shoemaker & Ramalho de Oliveira, 2008). It is vital for caregivers to recognize and understand patient experiences with their medications, as this awareness can foster a deeper connection and rapport between caregivers and patients (Shoemaker & Ramalho de Oliveira, 2008). Patients who face a higher medication burden—those who require daily medication—may elicit a more treatment-oriented perspective from caregivers. Furthermore, when caregivers engage in regular interactions with patients to administer medications, they create numerous opportunities to respond to alarms from HR monitoring tools, potentially enhancing the utilization of these devices within care practices.

## Methods

The main object of this project is to identify contextual and patient-level factors associated with utilization of the “Understand me!” heart rate-informed communication tool (Kildal et al., 2023) during a clinical testing phase of 2-3 months, where utilization is quantified primarily as hours of device use and secondarily as the caregiver’s response rate to system alarms and the number of registered situations.

### Hypotheses

#### Hypotheses 1

Patients in larger staffed residential units (≥ 8 staff) will have a lower proportion of alarms answered by caregivers (answered alarms/total alarms) than smaller units (< 8 staff), adjusted for patient function level (severity of ID, CP or level of care needed for somatic condition).

#### Hypotheses 2

There will be a positive correlation between the number of distinct caregivers and the proportion of alarms answered by caregivers (answered alarms/total alarms), adjusting for hours of device exposure and number of alarms.

#### Hypotheses 3

Patients classified as immobile (needs full mobility support/non-ambulatory) will have a higher proportion of alarms answered by caregivers (answered alarms/total alarms) than mobile patients, after accounting for hours of monitoring and number of alarms.

#### Hypotheses 4

Greater patient medication burden (operationalized as number of regular daily medications) will be associated with a higher proportion of alarms answered by caregivers (answered alarms/total alarms), compared with lower medication burden, after adjustment for patient function level (severity of ID, CP or level of care needed for somatic condition) and size of residential unit.

### Participants

The 40 study participants are included in an ongoing clinical trial by Kildal et al. (2023) (registered at ClinicalTrials.gov [NCT05738278]) (Kildal et al., 2023). Patients with communication difficulties are defined as having an impairment in the ability to convey concepts of both verbal and nonverbal form. The impairment is to such an extent that it causes caregivers to worry that the patient may experience pain and stress without being able to communicate it. Participants are recruited via Oslo University Hospital, Ahus Hospital, Oslo-, Bærum- and Drammen municipalities, which are located in south-eastern Norway.

Professional caregivers at the care homes will approach the parents or legal representatives of potential participants with written materials describing the study.

### Study design

The ongoing clinical trial that the current study is a part of includes a two-week *pre-intervention*, consisting of continuous HR registering across four potentially painful settings (e.g., physiotherapy); and an *intervention phase* consisting of formal change in procedure for the identified painful situation. The interventions are in one of four forms: changes in 1) physiotherapy techniques, 2) preparations for putting on casts, 3) lifting techniques or 4) personal hygiene procedures. The current study outcome measure is the caregiver response rate (proportion of alarms that were answered, e.g., 7 of 10). In addition, 1) activation level (number of days the tool has been activated during the usage period), 2) duration of use (total number of hours the tool has been in use), 3) level of detail in recording [how thoroughly the caregiver recorded context and observations (e.g., number of fields filled, free text use)] will be noted.

### Data collection

The data used in this study is collected across several sites in Oslo and the surrounding area with interventions administered at care homes with round-the-clock staff or one-to-one staffed school/daycare. This study will use already gathered data from Kildal et al. (2023) study, in addition to new data gathered.

Polar OH1 or Verity Sense armbands are used to calculate measures of HR. The current study data gathering will be using an armband that detects pulse at the upper arm (brachial artery), which is transmitted to an app developed for this study. The app is developed by the University Center for Information Technology (USIT) at the University of Oslo.

**Figure 1:**
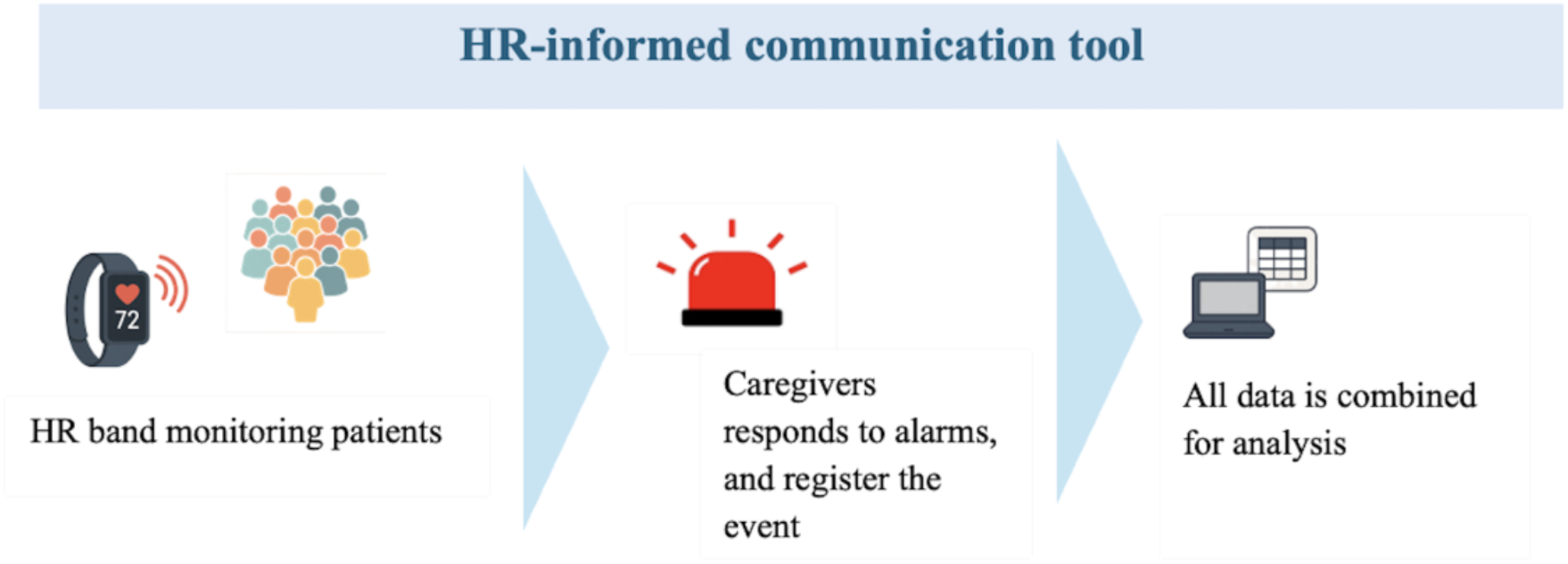
Overview of design. HR: Heart rate

### Analyses

To identify potential contextual and patient-level factors that may influence the utilization of the communication tool, we will compare the proportion of alarms answered by caregivers. To evaluate if the larger sized residential units have a higher proportion of alarms answered, descriptive statistics will be used. Pearson’s or Spearman’s, depending on whether the data is normally distributed or not, will be used to explore the possible correlation between the number of distinct caregivers and proportion of alarms answered. Multiple regression analysis or correlation analysis will be used to explore whether immobile patients will have a higher proportion of alarms answered than mobile patients, and to explore if greater medication burden will be associated with higher number of alarms answered, compared with lower medication burden.

### Inclusion criteria

Females and males between age 5 – 70 years of age are eligible to be included. All participants have PIMD and are MCP. All participants are evaluated with the Social Communication Questionnaire (Chandler et al., 2007) as part of evaluation of autism traits. Scores will be reported in the publication of results. Participants may have cerebral palsy to a degree that compounds the patient’s communication problems. Written informed consent must be obtained from the participant’s legal representative.

### Exclusion criteria

Exclusion criteria include not living in a care home with round-the-clock staff for at least five days a week; or not attending one-to-one staffed school/day-care at least five days a week.

Having any ongoing infection with a C-reactive protein (CRP) > 20 at the start of the data collection, or any type of cancer with ongoing chemotherapy are additional exclusion criteria

### Ethical considerations

To maintain confidentiality; evaluation forms, reports and other records will be identified by a coded number and initials only. All study records will be stored securely. The study has ethical approval from REK (2016/1956) and is registered at ClinicalTrials.gov [NCT05738278]. The study is conducted in accordance with the Declaration of Helsinki (World Medical Association, 2001).

## Discussion

The aim of this study is to increase knowledge of different contextual and patient-level factors that are associated with utilization of the “Understand me!” HR-informed communication tool. We will explore if larger size residential units affect the utilization of the HR-informed tool; see if higher number of distinct caregivers will have a lower response rate on alarms; explore if the patients mobility affects response rate on the alarms; and assess if medication use will be associated with a higher response rate of the alarms. The study require ingoing and thorough ethical awareness due to the patient’s inability to provide informed consent. Each caregiver will enter the participant into the trial, and researchers must be acutely aware of the MCPs signals of refusing participation. Altogether, this study will contribute to a better understanding of how various factors influence the utilization of HR-informed communication tools in the care of patients with PIMD, potentially leading to improvements in treatment practices and communication between patients and caregivers.

## Data Availability

Anonymised data produced in the present study are available upon reasonable request to the authors

## Abbreviations

PIMD: profound intellectual and multiple disabilities
MCP: minimally communicative patients
HR: heart rate

## Data availability

Not applicable. As this is a preregistration protocol, no data have been collected or analyzed yet.

## Author information

Department of Psychology, University of Oslo, Oslo, Norway

Maiken E. T. Willadsen & Daniel Quintana

Department of Neurohabilitation, Oslo University Hospital, Oslo, Norway

Bjørnar Hassel & Emilie S. M. Kildal

Department of Clinical Medicine, University of Oslo, Oslo, Norway

Bjørnar Hassel & Emilie S. M. Kildal

K.G. Jebsen, Centre for Neurodevelopmental Disorders, University of Oslo, Oslo, Norway

Daniel Quintana & Emilie S. M. Kildal

NevSom, Department of Rare Disorders, Oslo University Hospital, Oslo, Norway

Daniel Quintana

